# A Pilot Study on Cost and Other Implementation Factors Comparing Telehealth and In-Person Therapy Service Delivery Following NICU Discharge

**DOI:** 10.1101/2025.06.09.25329300

**Authors:** Roberta Pineda, Polly Kellner, Stacey Dusing, Cameron Kaplan, Rachel Chapman, Carol J. Peden

**Affiliations:** Chan Division of Occupational Science and Occupational Therapy, University of Southern California, Los Angeles, CA, USA; Keck School of Medicine, Department of Pediatrics, Los Angeles, CA, USA; Gehr Family Center for Health Systems Science and Innovation, University of Southern California, Los Angeles, CA, USA; Fetal and Neonatal Institute, Children’s Hospital of Los Angeles, Los Angeles, CA, USA; Division of Biokinesiology and Physical Therapy, University of Southern California, Los Angeles, CA, USA; Keck School of Medicine USC, Department of Anesthesiology, Los Angeles, CA, USA; University of Pennsylvania, Perelman School of Medicine, Department of Anesthesiology, Los Angeles, CA, USA

## Abstract

**Introduction:** Innovative models are needed to improve access to early therapy for high-risk infants discharged from the NICU. This study aimed to 1) compare costs between in-person and telehealth early therapy, and 2) evaluate adoptability, feasibility, adaptations, and acceptability of each model.

**Methods and Materials:** Twenty high-risk NICU infants were enrolled before discharge and randomized to receive therapy via telehealth or in-person Baby Bridge programming until community-based services began. Weekly visits were scheduled, with flexibility for switching formats (telehealth or in-person) when agreed upon by the therapist and family. Cost, utilization, and adaptations were tracked. Parent satisfaction was assessed via a post-discharge questionnaire.

**Results:** One infant was withdrawn due to readmission prior to receiving Baby Bridge services. Completion rates were high (18/19, 95%). In-person sessions were significantly more expensive ($141.35 ± $51.10) than telehealth sessions ($46.29 ± $16.19; p<0.001). Telehealth sessions generated positive average net revenue ($61.45 ± $54.31), while in-person sessions incurred losses (-$44.96 ± $63.63; p<0.001). Medicaid-insured sessions incurred losses for both telehealth (-$10.36 ± 4.94) and in-person (-$85.56 ± 38.53), whereas privately insured sessions yielded positive net revenues for telehealth ($91.92 ± 32.35) and in-person ($5.78 ± 51.20) sessions. No group differences were found in time to first session or session frequency. Visit format adaptations occurred in both groups (17% of telehealth visits; 36% of in-person; p<0.03). Parent satisfaction was comparable across groups.

**Conclusion:** Telehealth therapy is significantly less costly and more financially sustainable than in-person therapy. Both delivery models were feasible, with high satisfaction reported by families. Flexibility in adapting visit format supports better access and uptake, especially within in-person models. Telehealth offers a promising alternative for early intervention in high-risk infants following NICU discharge.

## Introduction

Neurodevelopmental impairment is already present at the time of discharge from the neonatal intensive care unit (NICU) (1–3) for many high-risk infants. Infants with identified neurodevelopmental impairment are at risk of long-term disability and often receive a referral for physical therapy, occupational therapy and/or speech-language pathology interventions to be initiated after NICU discharge (4, 5). Rapid changes in neurodevelopment continue after discharge, particularly during the first year of life (6, 7). This critical period presents an opportunity to positively impact outcomes through the provision of early and consistent therapy services (8, 9).

There is evidence supporting the use of early therapy interventions post-NICU discharge, with research identifying a positive impact on developmental skill acquisition and a reduction in the impact of abnormal postural reactions and asymmetries (10–27). Early therapy also enables clinicians to educate families, ensure adequate environmental and sensory exposures, improve oral feeding experiences, and address the psychosocial needs of the infant and family. Despite evidence supporting the benefits of early therapy services and policies requiring each state to offer early intervention (28), these services are often difficult to access (29–31). Many children with identified neurodevelopmental impairment prior to NICU discharge fail to get the therapy they need (30, 31), a problem that is further exacerbated for infants with social challenges (16).

The difficult transition from NICU to home further complicates early access to care (32, 33). We identified that preterm infants with neurodevelopmental impairment (n=57) in Missouri who were referred for therapy at the time of NICU discharge waited an average of 4 to 5 months before they received services, and many did not access any therapy (4). A preliminary study of access to early intervention in Los Angeles identified that less than half of infants referred for early therapy after NICU discharge were receiving services by 6 months of age (34). Missing the opportunity to impact function during this period of rapid brain development can stress families, increase societal burden, result in higher short- and long-term healthcare costs, and lead to a poorer transition to the educational system as the infants reach school age. Our team has identified this need and implemented a model of care, the Baby Bridge program, which is designed to improve access to early therapy services for at-risk infants in the first months after discharge.

The Baby Bridge program is an evidence-based implementation strategy aimed at enhancing early and continuous therapy services following NICU discharge for infants with alterations in neurodevelopment. The Baby Bridge program utilizes an occupational or physical therapist to see the infant and family in the NICU prior to discharge, conduct a comprehensive neurodevelopmental and feeding assessment to inform targeted interventions, and provide early therapy services in the home environment within one week of discharge and weekly thereafter, until other community-based services begin. While most of the program occurs after NICU discharge, a hallmark of the Baby Bridge program is that families establish rapport with the therapist while the infant is still in the NICU. This can occur through the therapist conducting the evaluation while the baby is still in the NICU, visiting with the family face-to-face, or through communications via phone, text, or email. The therapist also educates the family on ways to support their infant’s development between sessions, and offers support and assistance during the transition from hospital to home. The Baby Bridge therapist also functions as a navigator, assisting and guiding the family in receiving appropriate medical services. The Baby Bridge program was developed using stakeholder engagement, including interviews with health care professionals, parents, and early intervention leadership. The Baby Bridge program was implemented in Missouri and was shown to be related to faster initiation of therapy services (by an average of 89 days) from NICU discharge to the first therapy visit (35). The program was also shown to be sustainable after a 16-month implementation period, when billed to payors, which largely consisted of Medicaid (36).

Previous work on the Baby Bridge program investigated the in-person model. However, telehealth is now a widely accepted form of medical, therapy, and mental health service delivery (37–40), increasing the need for research on its cost, efficacy, and acceptability (41). The benefits of telehealth are variable, and more studies are emerging (41–43). Recent studies found telehealth can improve access to care and increase health care provider productivity (44–47). Significant cost savings can be realized when a telehealth model reduces provider drive time, particularly for families who live in rural areas (48–50). Telehealth services for neonates have been investigated by other disciplines (38, 51), but there is little evidence on telehealth therapy for this unique population. One study found that telehealth options increased access to care among safety net populations (44). In addition to the necessity of telehealth models for infection control due to the pandemic, telehealth models may also increase long term access especially in communities with large geographic areas to cover, complicated traffic patterns, and potentially limited availability of therapists to meet therapy needs across such areas.

To determine whether the same problems with access to early therapy services exist in Los Angeles, we reviewed the medical records of 79 preterm infants born at less than 32 weeks gestation who attended a 4- to 8-month corrected age visit at a high-risk infant follow up clinic. All infants had referrals for the state-funded early intervention program at NICU discharge. Additional referrals may have been made to outpatient services when wait times were extensive and/or there were significant, complex therapy needs. Less than half of these infants were receiving any therapy services 6 months after discharge (34).

As a component of a 3-part project funded by the American Occupational Therapy Foundation, the Baby Bridge model was adapted to a telehealth model following qualitative interviews with health care professionals who serve the therapy needs of infants and families following NICU discharge. In this previous study by the research team, telehealth Baby Bridge services were perceived to be an acceptable form of delivering early therapy services; participants recommended attempting at least one in-person visit to the NICU to build rapport; and suggestions to enhance telehealth were made (including supplementing telehealth sessions with occasional in-person sessions, using dolls or videos to demonstrate positioning and techniques, and having parents share videos of their infant between sessions to ensure more comprehensive feedback) (52).

Subsequently, the telehealth Baby Bridge model was defined to have these key pillars: 1) The therapist who would see the infant after discharge attempting an in-person session in the NICU prior to discharge to establish rapport and conduct assessments, 2) Setting up the first Baby Bridge session within 1 week of NICU discharge, 3) Providing at least weekly therapy sessions via telehealth until other community based therapy services commenced, and 4) offering flexibility in having an in-person session when deemed necessary by the family or treating therapist in order to optimize the therapy plan.

The telehealth model was tested for feasibility in 8 infants who received a referral for therapy at NICU discharge. The first telehealth session after NICU discharge occurred at an average of 6.0 ± 2.6 days following discharge. Infants received an average of 8.3 ± 2.1 telehealth sessions over 9.2 ± 3.5 weeks. All therapy sessions after NICU discharge were accomplished with telehealth rather than in-person sessions, and all families indicated they were “very satisfied” with Baby Bridge telehealth services.

This third component of the AOTF-funded work is to conduct a pilot study to assess differences in cost across in-person and telehealth early therapy sessions (specifically of the Baby Bridge program) in addition to assessing differences in adoptability, feasibility, adaptations, and acceptability.

## Methods

### Participants and setting

Participants included 20 high-risk parent-infant dyads hospitalized in an urban, level IV NICU. Consecutively eligible dyads from May 2023 to April 2024 were recruited if the infant was hospitalized in the NICU for more than 7 days; received a referral for post-NICU physical therapy, occupational therapy, and/or speech-language pathology (or for the state-wide early intervention program) at NICU discharge; was referred to the Baby Bridge program at least 48 hours before NICU discharge; had a chronological age of less than 6 months at the time of NICU discharge; and were from a family who spoke English. Infants who lived any distance away from the hospital were enrolled; however, infants who were not going to reside in the state of California following NICU discharge were excluded. Infants were deemed ‘high-risk’ for developmental challenges if they were recommended by the NICU medical team for therapy. Although we acknowledge that the term “high risk” can refer to high risk of medical complications or high risk of cerebral palsy, we use the term “high risk” to define the infant’s need for therapeutic interventions across a multitude of constructs including for language, cognition, self-regulation, sensory, motor, and feeding. A member of the research team attended weekly discharge rounds and reviewed the electronic medical record. Within discharge rounds, communication between the medical team and research team occurred, and the medical team referred infants to the Baby Bridge program through an online referral process in REDCap. The research team then screened and recruited those who were eligible.

Enrolled infants and families were randomized in REDCap using simple randomization to receive Baby Bridge services either through telehealth or in-person sessions. Parent-infant dyads were enrolled at least 2 days before NICU discharge to enable rapport to be established (between the Baby Bridge therapist and family) through a visit to the NICU when possible and via telephone, text or email messaging when an in-person visit was not possible prior to discharge. This first contact was attempted to be in-person in the NICU for both the telehealth and in-person groups. Enrolled infants and families then had a Baby Bridge telehealth or in-person session scheduled within one week of discharge (depending on their group assignment). Subsequent individual weekly sessions were adapted to in-person or telehealth (varying from assigned group) when needed or deemed appropriate (requested by the family or therapist or when an in-person session was felt to improve the quality of the therapy session). Weekly Baby Bridge programming was conducted until other therapy through the state-wide early intervention program commenced, meaning the infant was receiving therapy services as recommended. We also tracked rates of enrollment, rates and reasons for any cancellations, and completion of the program (defined as being seen until community-based early intervention services commenced). Infants were withdrawn if they were transferred to another hospital or if they were readmitted to the hospital and additional contact was not possible or feasible.

### Sociodemographic and medical data

Sociodemographic and medical data were collected from the electronic medical record for each dyad enrolled to better understand differences between groups as well as to define sample characteristics. Sociodemographic factors collected included: infant race, insurance type (public or private), maternal age, number of siblings, home distance from the hospital, and categorization of home residence (urban with more than 3,000 people per square mile, suburban with between 1,000 and 3,000 people per square mile, or rural with less than 1,000 people per square mile). Medical factors collected included: estimated gestational age at birth, the primary condition of the infant (congenital anomaly, preterm birth, or neurological condition not related to preterm birth or congenital anomaly), number of days of endotracheal intubation, number of days of hospitalization, and whether the infant was orally feeding at time of hospital discharge.

### In-person group assignment

For those assigned to the in-person Baby Bridge group, the first session was scheduled in the home within one week of NICU discharge, and attempts were made to see the infant weekly in the home environment for a one-hour therapy session. Scheduled sessions were confirmed with families via text messaging the day before each session. Sessions were rescheduled to telehealth in the event of therapist illness, parent preference, parent or child illness, or due to distance or therapist schedule limitations. Session adaptations were tracked along with their reasoning.

### Telehealth group assignment

For those assigned to the telehealth Baby Bridge group, a first telehealth session was scheduled within one week of discharge, and weekly telehealth sessions were scheduled thereafter. However, parents were informed that in-person sessions were an option should the need arise. Adaptations from a telehealth session to an in-person session were made in the event of therapist clinical judgment due to the medical complexity or feeding/tonal abnormalities of the infant and parent preference or rapport building. Session adaptations were tracked along with their reasoning.

### Implementation outcomes

We defined measures for this study based on previous implementation research (53–55). The primary outcome of interest for this study was the cost difference between telehealth and in-person sessions of the Baby Bridge program. We also investigated implementation outcomes of adoptability, feasibility, adaptations, and acceptability.

#### Cost

Cost is an important factor when determining feasibility of an intervention. When resources are limited, it can aid in choosing one intervention over another. For the purposes of this project, we estimated costs associated with each Baby Bridge session from a societal perspective. We did not assess cost by group assignment, due to the allowed adaptations to each session type, but instead pooled data from all in-person sessions and telehealth sessions. Cost was defined in relation to the organizational cost of providing Baby Bridge sessions and included the time that the therapist spent conducting therapy sessions (billable time), administrative time (documentation, communication with families), driving (for in-person sessions), and mileage reimbursement. The total billable and non-billable time that the therapist spent (including therapy session time, administrative time, and drive time) was multiplied by $50 per hour, which is the mean hourly wage for Occupational Therapists in California and the United States according to the 2023 Occupational and Employment and Wage Statistics from the Bureau of Labor Statistics (56). Mileage reimbursement was calculated at $0.67 per mile, the California mileage reimbursement rate for employees (57). Further, the number of billable minutes (in evaluation, for therapeutic activities, or therapeutic exercises) was contextualized using Healthcare Common Procedure Coding System (HCPCS) codes, and reimbursement amounts were estimated based on the current Fee For Service Medi-Cal (California Medicaid) payment schedule or approximated based on private insurance reimbursement rates for the CPT codes that would have been billed at the study site where this research took place. Due to variation in private insurance reimbursement rates, we took an average of the four most common private insurers at the study site to define the reimbursement potential. This enabled a total amount of reimbursement potential in context of the total cost of programming, with the ability to explore differences across telehealth and in-person costs, as well as Medicaid and private insurance reimbursement patterns.

#### Adoptability

Adoption refers to the uptake of Baby Bridge programming, measured at the organizational and provider level (53). This can include health care professionals’ buy-in and support of the program that enables infants and families to receive Baby Bridge services. We tracked the number of infants referred, approached, and enrolled. Adoption was operationalized as the total number of infants approached compared to those enrolled (i.e., enrollment rate).

#### Feasibility

Feasibility is defined as the extent to which Baby Bridge programming can be successfully used or carried out within a given agency or setting (53). To understand the feasibility of Baby Bridge telehealth programming, we captured timing (whether the first session was conducted within one week of discharge); average time from NICU discharge to first Baby Bridge session); frequency of sessions; rate and reasons for any therapy session cancellations; the completion rates (the percentage of enrolled infants who completed the program); and the timing of transition to community-based therapy. Feasibility was defined a priori as: the first Baby Bridge session being attempted within the first week; infants seen at least 3 sessions per month while in the program; and 80% of participants successfully transitioning to early intervention programming.

#### Adaptations

Adaptations include intentional changes made to programming in order to enable real-world uptake by aligning the intervention to the context in which it exists (58). To aid in understanding specific adaptations made to telehealth versus in-person programming, we captured the proportion of sessions conducted via telehealth within the telehealth group and proportion of sessions that occurred in-person within the in-person group. Reasoning for such adaptations was tracked as described above in the telehealth and in-person assignment sections.

#### Acceptability

Acceptability is the perception among stakeholders, specifically the consumers or parents, that the Baby Bridge program is satisfactory. Using Proctor’s model (53) as a guide and through adaptation of a previously reported measure of acceptability (59), the Baby Bridge Parent Survey was developed to determine if programming was acceptable to parents. The survey was developed, and five members of the research team added/revised items to aid in comprehensive collection of perceptions as well as to aid clarity of each question’s wording. It went through an iterative process with multiple revisions until no further feedback from team members was received and there was consensus on the inclusion and wording of survey questions. The survey took less than 5 minutes to complete.

Upon completion of the Baby Bridge program, parents were asked to complete the Baby Bridge Parent Survey to gather their perceptions about Baby Bridge programming. Survey questions were loaded into REDCap and administered electronically through a shared link to a REDCap survey. Parents received subject remuneration of $50 after completing the survey. Parent satisfaction was measured based on a summed score (possible range of 0-10) of the following questions from the survey:

1. Were therapy goals achieved through Baby Bridge programming (2=yes, 1=partially, 0=no)
2. How likely would you be to recommend the Baby Bridge program to other NICU parents? (3=definitely will, 2=probably will, 1=probably will not, 0=definitely will not)
3. How likely would you be to use Baby Bridge programming again? (3=definitely will, 2=probably will, 1=probably will not, 0=definitely will not)
4. Was there anything about the Baby Bridge program that you found confusing or complicated? (0=yes, 1=no)
5. Did you experience any challenges with he Baby Bridge program (0=yes, 1=no).

### Analysis

IBM SPSS Statistics (version 28) was used for statistical analysis. A power analysis to determine sample size was not conducted, as this was a pilot study aimed at understanding initial differences across groups while determining variance to aid in future sample size determination for a larger trial. Sociodemographic and medical factors are reported descriptively and differences across groups were investigated using independent samples t-tests and chi-square analyses. Any missing data was excluded from data analysis.

#### Cost

The costs of in-person sessions vs. telehealth sessions are reported descriptively. Differences in costs across telehealth and in-person sessions were investigated using independent samples t-tests and linear regression models.

#### Adoptability

Rates of enrollment are reported descriptively but not analyzed between groups, as enrollment occurred prior to group assignment.

#### Feasibility

We investigated differences across groups (in-person versus telehealth) in whether the first session occurred within 1 week of NICU discharge, average number of days between NICU discharge until first Baby Bridge session, frequency of sessions, number of therapy session cancellations, and the completion rates using independent samples t-tests and chi-square analyses.

#### Adaptations

The proportion of sessions conducted within the assigned group was investigated across groups using independent samples t-tests.

#### Acceptability

Satisfaction scores were compared across groups using independent samples t-tests.

## Results

Thirty infants were referred for the Baby Bridge program by the medical team, of whom 26 were eligible. Twenty infants were enrolled (77%) between May 2023 and April 2024, with 10 randomized to telehealth (50%) and 10 randomized to in-person (50%) Baby Bridge services. One infant was withdrawn due to readmission prior to receiving Baby Bridge services. Eighteen of the 19 remaining infants completed Baby Bridge programing (95%); 1 became unresponsive to contact after NICU discharge and prior to Baby Bridge services being initiated. See Table 1 for sociodemographic and medical characteristics of the 18 participants who completed Baby Bridge programming. The telehealth and in-person groups did not differ in any of the documented characteristics.

**Table 1.**
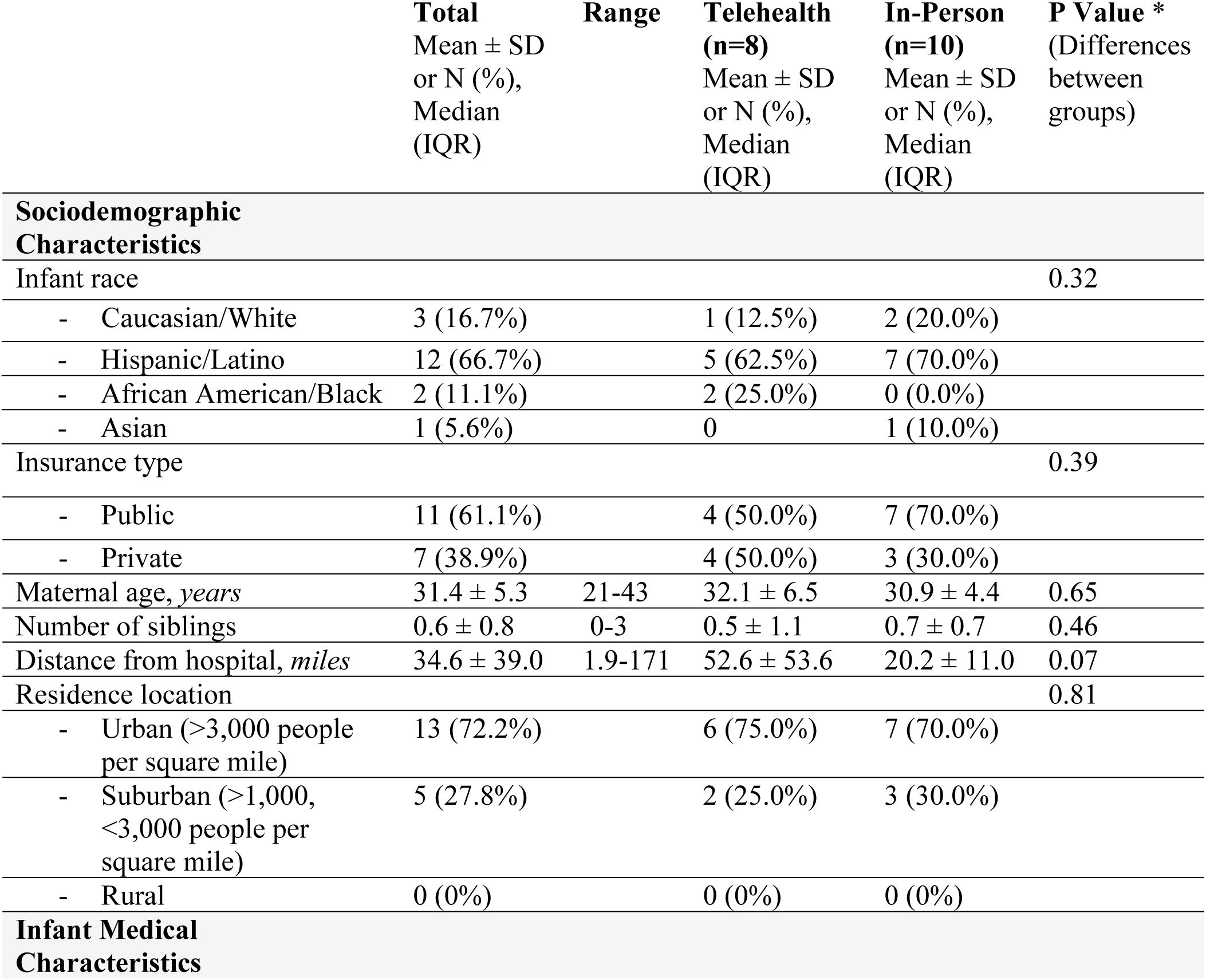
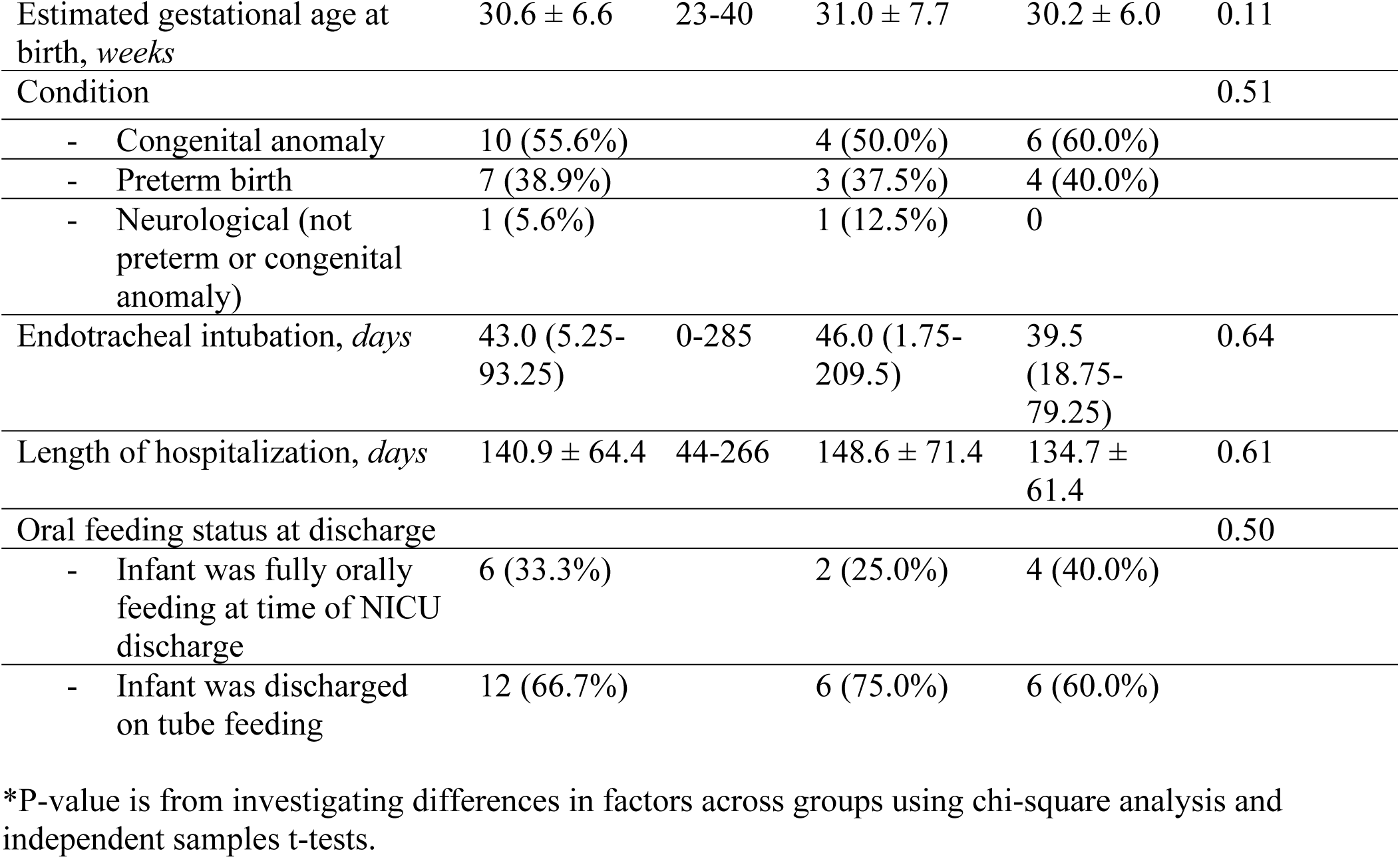
Maternal and social characteristics of the dyads who received the Baby Bridge telehealth or in-person programming (n=18)

### Cost

Cost analysis between telehealth vs. in-person Baby Bridge sessions is reported in Table 2. Average total cost of each in-person therapy session was higher ($141.35 ± $51.10) than a telehealth session ($46.29 ± $16.19; p<0.001), with average net revenue for telehealth sessions being higher ($61.45 ± $54.31) than the net losses for in-person sessions ($-44.96 ± $63.63; p<0.001). This was due to therapist drive time, mileage reimbursement, and the increased time the therapist spent with the family during in-person sessions.

**Table 2.**
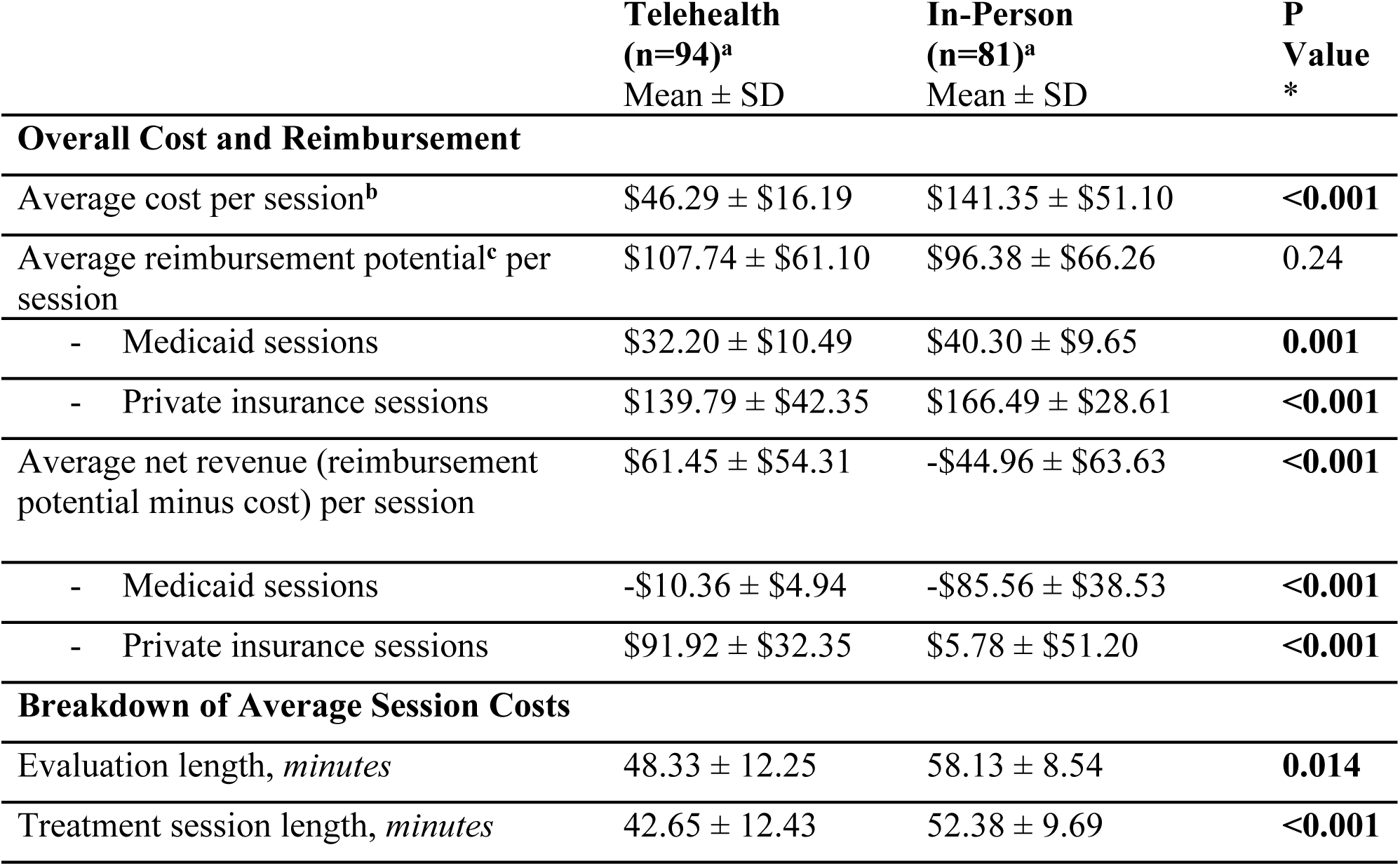
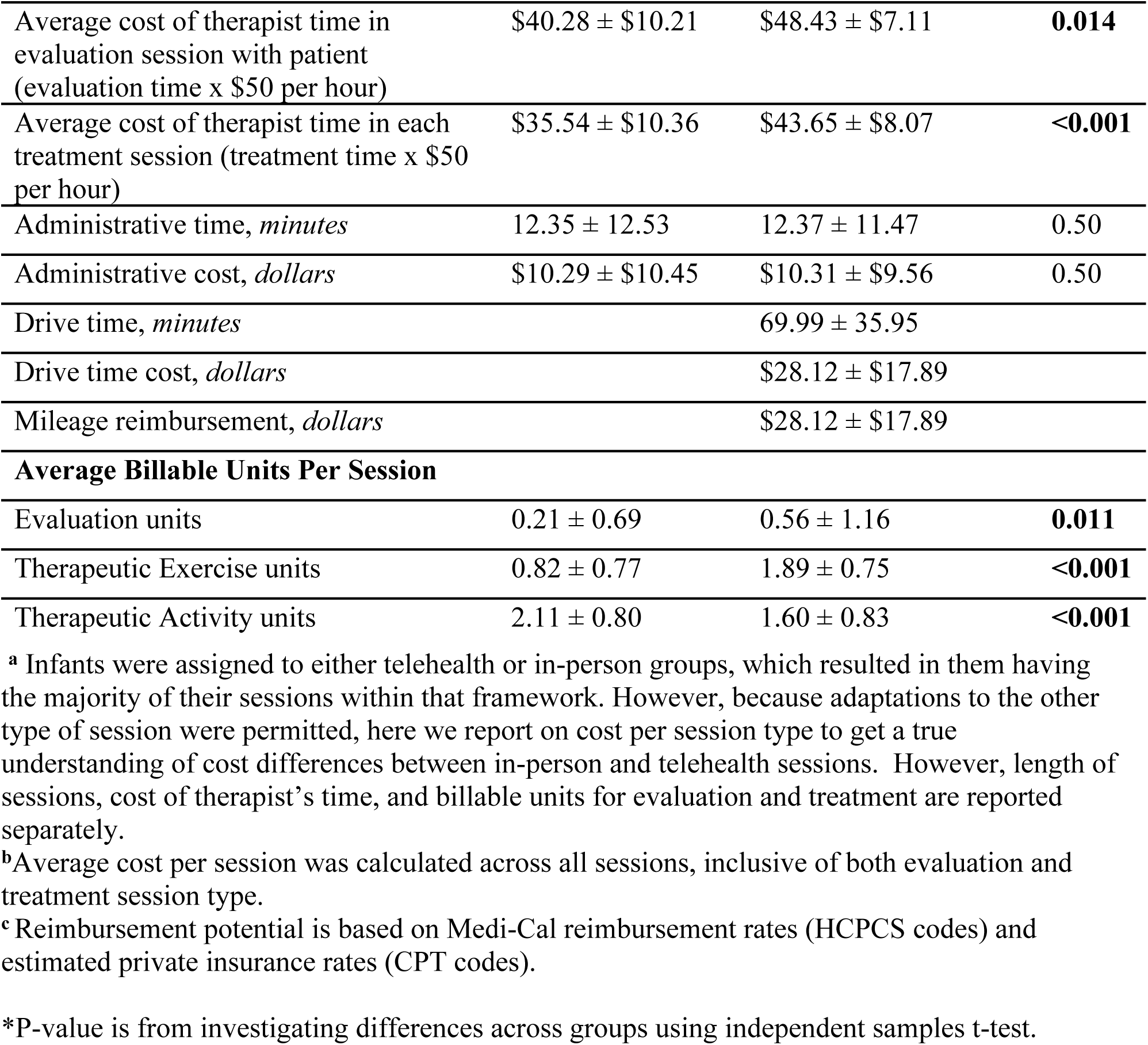
Cost analysis between telehealth and in-person sessions.

Average negative net revenues for Medicaid were -$10.36 ± 4.94 for telehealth and - $85.56 ± 38.53 for in-person sessions, compared to positive net revenues for private insurance of $91.92 ± 32.35 and $5.78 ± 51.20, respectively. With our sample’s payor mix of 60% public and 40% private, the program overall yielded a positive net revenue of $2,134.11 for all 175 telehealth and in-person sessions combined. However, the mix of 94 (53%) sessions delivered via telehealth and 81 (46%) delivered in-person also contributed to sustainability, as there was more total net revenue for all telehealth sessions ($3,642,47) compared to losses for all in-person sessions of -$1,508.36; p<0.001). Administrative time (documentation and communication with families) did not differ between groups. The therapeutic exercise CPT code was used more often during in-person sessions, an average of 1.89 units per session compared to 0.82 units per session for telehealth sessions (p<0.001). Conversely, the therapeutic activity code was used more frequently for telehealth sessions, an average of 2.11 units per session compared to 1.60 units per session for in-person sessions (p<0.001), which impacted the reimbursement potential for each session.

### Adoptability

See Figure 1 for a flow diagram of eligible and enrolled infants in the study. 30 infants were referred prior to NICU discharge by the NICU medical team, of which 26 were eligible. 4 of those dyads were referred but could not be reached prior to discharge. Of the 22 who were approached for consent, 20 (91%) enrolled in the program. The 2 dyads (9%) who did not enroll in the program asked for time to consider participation in the study after they were initially approached and then were unresponsive to further contact despite multiple methods of communication (phone call, text message, in-person approach in NICU) and at least 3 attempts.

**Figure 1.**
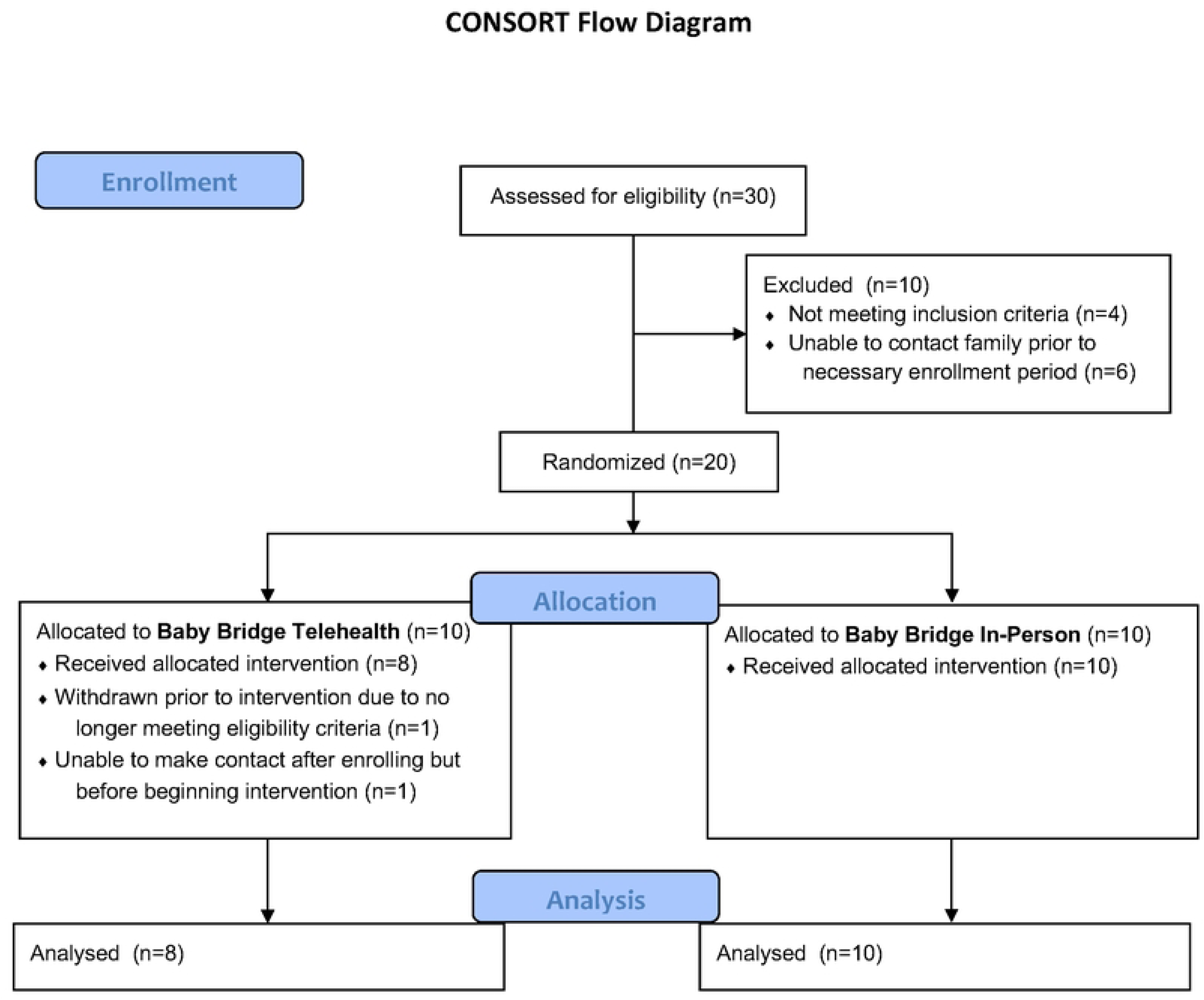
Flow diagram of participants.

Out of the 20 enrolled, 1 (5%) infant was withdrawn due to prolonged readmission with lack of continued engagement by the family. Of the 19 who remained in the study, 1 (5%) infant did not complete the program because they stopped responding to contact after NICU discharge. The maternal, sociodemographic, and infant medical factors of this dyad did not appear to differ from that of the rest of the study sample. The remaining 18 dyads (95%) received Baby Bridge services until community-based early intervention commenced.

Among the 18 infants who completed Baby Bridge programming, the average gestational age at birth was 30.6 ± 6.6 weeks. Diagnoses represented in the sample included congenital anomaly (56%), preterm birth (39%), and neurological diagnosis in absence of congenital anomaly or prematurity (6%). Evaluation for the Baby Bridge program occurred at an average of 50.0 ± 7.8 weeks postmenstrual age. The average gestational age at birth, insurance type (public or private), and racial breakdown of the sample recruited for the program were similar to that of other, unrelated samples for this study site (60).

### Feasibility

Findings related to feasibility of service uptake can be found in Table 3. Of the 18 dyads who received Baby Bridge services, the first session occurred between 3 and 21 days after discharge at an average of 6 ± 4 days following discharge. All (100%) participants were offered a session within 1 week of discharge, and 17 (94%) received their first session within one week of discharge. The one dyad that did not receive a session within 1 week of discharge had their first session rescheduled a few times due to family preference in the midst of daily medical appointments that required significant driving time for them in the weeks following discharge. There were no differences in timing to first therapy session based on group assignment. An in-person evaluation was attempted for all dyads, and 14 of the dyads (78%) ultimately received an in-person evaluation. Reasons why 4 did not receive an in-person evaluation were due to inadequate timing between referral and NICU discharge (n=2) as well as therapist availability and privileges for in-person evaluation in the hospital (n=2). These 4 had attempts at in-person evaluations in the home, but they were not possible due to distance from hospital (n=4).

**Table 3.** Service uptake in Baby Bridge telehealth and in-person groups (n=18)

| **Service Uptake** | **Total**<br>**Mean ± SD**<br>**or N (%),**<br>**Median**<br>**(IQR)** | **Range** | **Telehealth**<br>**(n=8)**<br>**Mean ± SD**<br>**or N (%),**<br>**Median**<br>**(IQR)** | **In-Person**<br>**(n=10)**<br>**Mean ± SD**<br>**or N (%),**<br>**Median**<br>**(IQR)** | **P**<br>**Value \*** |
| --- | --- | --- | --- | --- | --- |
| Postmenstrual age at the time of NICU discharge, *weeks* | 50 ± 7.8 | 38-64 | 51.4 ± 8.9 | 48.9 ± 7.2 | 0.54 |
| Time from NICU discharge to first Baby Bridge session, *days* | 6.0 ± 4.0 | 3-21 | 7.0 ± 5.6 | 5.0 ± 1.8 | 0.61 |
| Number of Baby Bridge sessions | 9.7 ± 3.4 | 4-20 | 8.3 ± 3.4 | 10.9 ± 4.2 | 0.09 |
| Length of time Baby Bridge sessions were completed, *weeks* | 11.9 ± 7.0 | 5.6-25.9 | 11.3 ± 4.9 | 14.7 ± 6.6 | 0.61 |
| Number of sessions per month | 3.4 ± 1.0 | 1.7-5.5 | 3.4 ± 1.2 | 3.5 ± 1.0 | 0.40 |
| Transitioned to early intervention services | 18 (100%) |  | 8 (100%) | 10 (100%) | 1.00 |
| Infant age at time of transition to early intervention, *corrected months* | 8.5 ± 2.7 | 3.9-10.6 | 7.5 ± 2.7 | 7.9 ± 2.9 | 0.85 |
| Average parent satisfaction score | 11.6 ± 0.6 | 10-12 | 11.6 ± 0.7 | 11.6 ± 0.5 | 0.57 |

\*P-value is from investigating differences across in-person and telehealth groups using independent samples t-test or Chi-square analysis.

Those infants who completed the program received an average of 9.7 ± 4.0 sessions over 11.9 ± 7.0 weeks, which did not differ across groups. On average, participants received 3.4 ± 1.0 sessions per month, which did not differ across groups. Reasons for missed sessions were patient no-shows (1% of scheduled sessions), patient cancellations (17% of scheduled sessions), and therapist cancellations (2% of scheduled sessions). Missed sessions due to patient no-shows or patient cancellations did not differ between the telehealth or in-person group. The 3 missed sessions due to therapist cancellations only occurred in the in-person group, and thus were significantly higher (p<0.001) in the in-person group. Among the 19 infants who were enrolled and remained in the study after discharge, 18 (95%) completed Baby Bridge programming as defined by receiving Baby Bridge sessions until community-based early intervention services commenced. There were no differences across groups in relation to timing to first Baby Bridge session, whether the infant transitioned to community-based services, and the age of the infant at time of that transition. No harms or unintended events were reported to the PI or Baby Bridge therapist.

Feasibility was achieved based on our a priori definitions with Baby Bridge sessions attempted within the first week for all enrolled infants, the average number of sessions per month being 3.4 ± 1.0, and 100% of infants successfully transitioning to early intervention programming.

### Adaptations

There were a total of 175 scheduled sessions across the sample. Dyads in the telehealth group had a total number of 66 scheduled sessions, and dyads in the in-person group had a total of 109 scheduled sessions. 50 (29%) of all sessions were adapted from their assigned (telehealth or in-person) group. 11 (17%) of telehealth group sessions were adapted to in-person, and 39 (36%) of in-person group sessions were adapted to telehealth. Significantly more sessions were adapted in the in-person group as compared to the telehealth group (p=0.03). Telehealth sessions were adapted to in-person due to medical complexity/feeding/tonal abnormalities (n=8, 12%) and parent preference/rapport building (n=6, 9%). In-person sessions were adapted to telehealth due to therapist illness (n=10, 9%), parent preference or parent/child illness (n=16, 15%), and distance or therapist schedule limitations (n=10, 9%).

### Acceptability

All 18 families who received any Baby Bridge sessions completed the Baby Bridge Parent Survey at the end of the program. Average parent satisfaction scores were high with an average of 11.6 ± 0.6 (out of a possible 12) and did not differ between the telehealth and in-person groups (p=0.57).

## Discussion

The key findings of this study are that the total cost was significantly less for telehealth sessions compared to in-person sessions. Sustainability could be achieved based on reimbursement potential for both telehealth and in-person service delivery among the privately insured, however, it would not be sustainable if all patients were covered by Medicaid. Evaluations and therapy sessions were longer in duration for in-person sessions, yet net revenue was higher for telehealth sessions due to therapist drive time and mileage reimbursement costs related to in-person sessions. The feasibility of conducting weekly therapy within one week of discharge and weekly thereafter was similar across in-person and telehealth models. Parent satisfaction was similar across the in-person and telehealth groups. Both types of assigned sessions were adapted based on need, with more adaptations of in-person sessions to telehealth sessions. Parent satisfaction was similar for families receiving in-person and telehealth early therapy sessions.

Cost is a driving factor related to implementation of programming that can impact human health. On average, each telehealth session was $95.06 less than each in-person session. In-person evaluation and treatment sessions were, on average, longer than telehealth sessions. In addition to the longer session time (and associated cost), in-person sessions incur the additional cost of drive time as well as mileage reimbursement. This can vary based on the catchment area for each therapist and across communities with different rural versus urban make-up and different traffic patterns. We observed that the average loss per session was much lower for telehealth sessions than for in-person sessions. Since this difference is mostly driven by differences in non-billable time (e.g. driving), the difference would likely remain consistent across settings with different payor mixes. However, losses for both session types may be significantly less when reimbursement is higher.

Reimbursement potential and sustainability for Baby Bridge programming, whether through an in-person model or via telehealth, depend on payor mix. Our revenue potential was estimated based on Medicaid and private insurance reimbursement rates. We found that such Baby Bridge programming would not be sustainable with 100% Medicaid reimbursement. Our sample yielded a positive net revenue with a ∼60% Medicaid and ∼40% private insured mix, but variations in payor mix will affect the sustainability of the program. Further, different states and locations may have different reimbursement rates that could change the reimbursement potential for both the privately and publicly insured.

This is the first study, to our knowledge, that has compared cost across telehealth and in-person sessions of early therapy. However, more research is needed to better understand the nuances of longer session time and how the service delivery type may impact quality and the comprehensiveness of each session. Further evidence that there may be differences in the actual therapy delivery in each session type (telehealth vs. in-person) is that there were significant differences in use of billable therapeutic exercise (targeting strength/range of motion) compared to therapeutic activities (targeting functional, real-world activities). Reduced opportunities for hands-on coaching of exercises may result in increased opportunities for coaching, explaining, and educating on activities as they relate to daily living. This can impact reimbursement potential for each session, but may also reflect differences in therapy approach across the two models.

The telehealth and in-person models appeared to have similar completion rates (90% compared to 100%), with one infant who was in the telehealth group not completing programming. However, it is important to note that not all infants who were eligible for programming were able to be enrolled. The process of enrollment by the research team was persistent and provided multiple opportunities for enrollment. As such, without such an initiative, special care may be needed to ensure families have access to the services they can benefit from. There was a high rate of completion among those who enrolled, with one infant being withdrawn due to rehospitalization and the other (who was assigned to the telehealth group) not being reachable after initial consent. It is unclear if this could be an indication that before initiating services, there was some parent preference for in-person services. This phenomenon has been backed up in the literature (61). Unfortunately, this family became unresponsive to contact, so this cannot be confirmed. Satisfaction rates were equally high in the telehealth and in-person groups, which is also reflected in the literature with other populations and types of healthcare programming (62, 63). This is also likely influenced by the fact that we ultimately offered a hybrid model, which allowed for in-person or telehealth sessions based on therapist/family needs and preferences.

Adaptations occurred across both the in-person and telehealth groups, with more changes to telehealth among those originally assigned to the in-person group. The decision to allow for adaptations to increase access to care and improve quality of services provided was made following a feasibility trial on Baby Bridge telehealth services (not yet published). While allowing adaptations makes interpretation across groups more nuanced, it appeared to be an critical factor in uptake. Telehealth services are criticized for not allowing full engagement with families, and, especially in the midst of significant medical complexity, the additional opportunities for observation, physical assessment, and rapport building were believed to be important to the therapist providing services. Likewise, when there were barriers to in-person sessions, such as transportation challenges or illness, telehealth enabled continuity of care for a population at high risk for transportation barriers and rehospitalization.

This study had several limitations. It was a small pilot study intended to lay the groundwork for future investigation. The hybrid model complicates comparison across models and limits conclusions about each delivery type. Not all eligible infants could be enrolled, introducing potential sample bias. Reasoning as to why parents did not respond to recruitment calls could not be ascertained, limiting our understanding as to why some did not enroll. The sample was recruited from a NICU with high medical complexity and long hospital stays, making it challenging to time recruitment with eventual discharge across many eligible infants. Goals were set with the family in both groups, irrespective of group assignment, but differences in actual therapy session content and how that aligned with goals were not explored. Nuanced differences in the type and quality of therapy provided was not ascertained in this small study but could provide important information in future studies.

## Conclusion

Telehealth sessions were observed to cost less than in-person sessions for early therapy after NICU discharge. However, adaptations appear to be an important component of care in both models. In-person and telehealth delivery models were similarly adoptable and feasible, with comparable levels of parent satisfaction. Future research should focus on understanding how therapy delivery differs between telehealth and in-person sessions, particularly in terms of quality and comprehensiveness.

## Data Availability

The data supporting this study's findings are available upon request from the corresponding author.

## Acknowledgements

We wish to thank Bryant Edwards, Cheryl Garden, Philipe Friedlich, Cynthia Gong, Sahar Ghahremani, Ariel Galan, Jennifer Koo, Yang Li, Camilla Catignas, Bethany Gruskin, Sharon Han, Maquela Noel, Marinthea Richter, Tara Crapnell, Maya Misikoff, and Carolyn Ibrahim.

